## Supplementary Table 1 for "Evaluation of the safety and immunogenicity of fractional intradermal COVID-19 vaccines as a booster: A pilot study"

**Supplementary Table 1.** Baseline characteristics of the subjects of each group in initial phase.

| Primary series (IM) - Booster (ID) | | Types of vaccines | | | | | | | |
|---|---|---|---|---|---|---|---|---|---|
| | | All | CoronaVac - BNT162b2 | CoronaVac - ChAdOx1 | ChAdOx1 - BNT162b2 | ChAdOx1 - ChAdOx1 | BNT162b2 - BNT162b2 | BNT162b2 - ChAdOx1 | *p*-value |
| **Initial phase** | | | | | | | | | |
| Number of subjects | n<br>% | 58<br>100.00 | 10<br>17.24 | 10<br>17.24 | 10<br>17.24 | 10<br>17.24 | 9<br>15.52 | 9<br>15.52 | - |
| Age (years) | Median (IQR) | 43.00 (35.00, 48.00) | 40.50 (28.00, 45.00) | 42.00 (24.00, 44.00) | 49.00 (46.00, 51.00) | 45.00 (43.00, 50.00) | 41.00 (36.00, 43.00) | 39.00 (35.00, 43.00) | 0.323 |
| Male | n<br>% | 19<br>(32.76) | 3<br>15.79 | 3<br>15.79 | 2<br>10.53 | 1<br>5.26 | 7<br>36.84 | 3<br>15.79 | 0.055 |
| Body mass index: BMI ($kg/m^2$) | Median (IQR) | 23.40 (20.60, 25.90) | 25.55 (24.40, 27.50) | 20.00 (18.00, 23.20) | 24.65 (22.30, 25.90) | 22.40 (20.80, 27.10) | 24.10 (20.70, 24.50) | 22.60 (20.90, 23.60) | 0.181 |
