## Supplementary Table 2 for "Evaluation of the safety and immunogenicity of fractional intradermal COVID-19 vaccines as a booster: A pilot study"

**Supplementary Table 2.** Immunogenicity response following intradermal booster vaccination by types of vaccines in primary series (2-dose intramuscular) and booster (ID) in the initial phase.

| Primary series (IM) - Booster (ID) | Types of vaccines | | | | | | | |
|---|---|---|---|---|---|---|---|---|
| | **All** | **CoronaVac - BNT162b2** | **CoronaVac - ChAdOx1** | **ChAdOx1 - BNT162b2** | **ChAdOx1 - ChAdOx1** | **BNT162b2 - BNT162b2** | **BNT162b2 - ChAdOx1** | ***p*-value** |
| **Number of subjects** | **(n=58)** | **n=10** | **n=10** | **n=10** | **n=10** | **n=9** | **n=9** | |
| **anti-RBD IgG (BAU/mL)** | | | | | | | | |
| GMC at pre-booster vaccination (95% CI) | 218.66 (168.09, 284.44) | 94.87 (51.88, 173.49) | 112.93 (65.94, 193.39) | 188.60 (119.77, 296.97) | 240.80 (126.88, 457.00) | 353.21 (200.27, 622.96) | 755.29 (479.91, 1188.69) | <0.001* |
| *p*-value | | 0.632 | | 0.491 | | 0.028* | | |
| GMC at post-booster vaccination (95% CI) | 1,188.24 (889.78, 1586.81) | 3,209.64 (1974.92, 5216.32) | 2,810.93 (1850.26, 4270.39) | 1,489.54 (797.55, 2781.92) | 309.48 (179.16, 534.62) | 848.58 (413.62, 1740.94) | 734.93 (437.25, 1235.29) | <0.001* |
| *p*-value | | 0.645 | | <0.001* | | 0.713 | | |
| GMR between post-booster vaccination /pre-booster vaccination (95% CI) | 5.43 (3.66, 8.06) | 33.83 (19.34, 59.16) | 24.89 (14.14, 43.81) | 7.90 (5.80, 10.76) | 1.29 (1.10, 1.50) | 2.40 (1.52, 3.81) | 0.97 (0.80, 1.19) | <0.001* |
| *p*-value | | 0.394 | | <0.001* | | <0.001* | | |
| **Live virus plaque reduction neutralization titers ($PRNT_{50}$) post intradermal boosting** | | | | | | | | |
| GMT against delta strain at post-booster vaccination (95% CI) | 200.64 (154.07, 261.29) | 409.40 (257.48, 650.96) | 431.83 (258.86, 720.36) | 300.78 (184.08, 491.46) | 95.60 (58.82, 155.37) | 116.52 (52.38, 259.21) | 96.99 (48.31, 194.72) | <0.001* |
| *p*-value | | 0.863 | | 0.002* | | 0.695 | | |
| GMT against omicron strain at post-booster vaccination (95% CI) | 27.52 (14.57, 51.97) | 76.34 (39.57, 147.29) | 130.21 (72.30, 234.51) | 51.75 (24.89, 107.57) | 6.71 (1.29, 34.74) | 53.31 (26.87, 105.78) | 1.94 (0.09, 43.11) | <0.001* |
| *p*-value | | 0.188 | | 0.019* | | 0.029* | | |
| Number (%) with $PRNT_{50}$ against omicron strain $\leq$1:10 | 6 (10.34) | - | - | - | 3 (30.00) | - | 3 (33.33) | 0.017* |

Notes: 1) * $p \leq 0.05$
2) One-way ANOVA and unpaired t-test with parametric assumptions satisfied was determined *P*-value among those who received any vaccinations

3) Abbreviation: BAU/mL: binding antibody unit/mL, GMC: geometric mean concentration, GMT: geometric mean titer, GM: geometric mean, GMR: geometric mean ratio
