## Supplementary Table 3 for "Evaluation of the safety and immunogenicity of fractional intradermal COVID-19 vaccines as a booster: A pilot study"

**Supplementary Table 3.** Immunogenicity response following intradermal booster vaccination by types of vaccines in primary series (2-dose intramuscular) and booster (ID) in the extended phase.

| **Primary series (IM) - Booster (ID)** | **Types of vaccines** | | | | |
|---|---|---|---|---|---|
| | **All** | **CoronaVac - BNT162b2** | **CoronaVac - ChAdOx1** | **ChAdOx1 - BNT162b2** | ***p*-value** |
| **Number of subjects** | **(n=135)** | **n=45** | **n=45** | **n=45** | |
| **anti-RBD IgG (BAU/mL)** | | | | | |
| GMC at pre-booster vaccination (95% CI) | 54.13 (42.99, 68.15) | 34.26 (23.76, 49.39) | 27.52 (18.71, 40.46) | 168.21 (137.42, 205.90) | <0.001* |
| GMC at post-booster vaccination (95% CI) | 1,494.31 (1308.69, 1706.27) | 1,978.48 (1595.09, 2454.02) | 1,326.85 (1025.61, 1716.57) | 1,271.08 (1032.54, 1564.71) | 0.011 |
| GMR between post-booster vaccination /pre-booster vaccination (95% CI) | 27.61 (21.98, 34.68) | 57.75 (42.53, 78.42) | 48.22 (33.40, 69.62) | 7.56 (6.34, 9.00) | <0.001* |
| **Live virus plaque reduction neutralization titers ($PRNT_{50}$) post intradermal boosting** | | | | | |
| GMT against delta strain at post-booster vaccination (95% CI) | 257.20 (221.85, 298.18) | 287.62 (225.33, 367.14) | 206.15 (149.94, 283.41) | 286.95 (235.14, 350.19) | 0.112 |
| GMT against omicron strain at post-booster vaccination (95% CI) | 45.52 (38.24, 54.19) | 52.74 (38.10, 72.99) | 45.82 (32.98, 63.66) | 39.05 (29.96, 50.89) | 0.382 |
| GMR: delta/omicron at post-booster vaccination (95%CI) | 5.65 (4.89, 6.52) | 5.45 (4.11, 7.24) | 4.50 (3.53, 5.73) | 7.35 (5.94, 9.10) | 0.020* |
| Number (%) with $PRNT_{50}$ against omicron strain ≤1:10 | 19 (9.63) | 6 (13.33) | 5 (11.11) | 2 (4.44) | 0.331 |
| **Number of subjects** | **(n=60)** | **n=20** | **n=20** | **n=20** | |
| **ELISpot responses (SFU/$10^6$ cells)** | | | | | |

| | | | | | |
|---|---|---|---|---|---|
| ELISpot-S<br>GM at pre-booster vaccination<br>(95%CI) | 11.01<br>(6.80,<br>17.81) | 4.22<br>(2.07,<br>8.61) | 5.89<br>(2.79,<br>12.41) | 53.68<br>(28.15,<br>102.37) | <0.001* |
| ELISpot-NMO<br>GM at pre-booster vaccination<br>(95%CI) | 8.31<br>(5.39,<br>12.82) | 9.41<br>(4.37,<br>20.27) | 20.15<br>(9.98,<br>40.68) | 3.03<br>(1.60,<br>5.75) | <0.001* |
| ELISpot-S<br>GM at post-booster vaccination<br>(95% CI) | 105.43<br>(71.88,<br>154.64) | 129.75<br>(62.58,<br>269.02) | 46.82<br>(25.16,<br>87.14) | 192.93<br>(108.44,<br>343.22) | 0.006* |
| ELISpot-NMO<br>GM at post-booster vaccination<br>(95% CI) | 25.24<br>(15.19,<br>41.93) | 45.97<br>(18.02,<br>117.28) | 21.35<br>(10.88,<br>41.88) | 16.38<br>(5.58,<br>48.14) | 0.229 |
| ELISpot-S<br>GMR: post-booster vaccination<br>/pre-booster vaccination (95% CI) | 9.58<br>(6.15,<br>14.91) | 30.77<br>(12.69, 74.62) | 7.95<br>(4.22,<br>14.97) | 3.59<br>(2.14,<br>6.03) | <0.001* |
| ELISpot-NMO<br>GMR: post-booster vaccination<br>/pre-booster vaccination (95% CI) | 3.04<br>(1.84,<br>5.02) | 4.88<br>(1.98,<br>12.04) | 1.06<br>(0.67,<br>1.67) | 5.41<br>(18.4,<br>15.89) | 0.010* |

Notes: 1) * $p \leq 0.05$
2) One-way ANOVA and was determined *p*-value among those who received any vaccinations
3) Abbreviation: BAU/mL: binding antibody unit/mL, GMC: geometric mean concentration, GMT: geometric mean titer, GM: geometric mean, GMR: geometric mean ratio, SFU/$10^6$ cells: spot forming unit per million cells
