## Supplementary Table 4 for "Evaluation of the safety and immunogenicity of fractional intradermal COVID-19 vaccines as a booster: A pilot study"

**Supplementary Table 4.** Adverse events following intradermal COVID-19 vaccination in the booster in the extended phase.

| Adverse events | Type of vaccines | | | | *p*-value |
|---|---|---|---|---|---|
| **Primary series (IM) - Booster (ID)** | **All** | **CoronaVac - BNT162b2** | **CoronaVac - ChAdOx1** | **ChAdOx1 - BNT162b2** | |
| **Number of subjects** | **n=135** | **n=45** | **n=45** | **n=45** | |
| **Injection site reaction, n (%)** | 122 (90.37) | 40 (88.89) | 41 (91.11) | 41 (91.11) | 0.148 |
| Mild, n (%) | 98 (72.59) | 31 (68.89) | 29 (64.44) | 38 (84.44) | |
| Moderate, n (%) | 24 (17.78 | 9 (20.00) | 12 (26.67) | 3 (6.67) | |
| **Any Systemic reaction, n (%)** | 107 (79.26) | 37 (82.22) | 39 (86.67) | 31 (68.88) | 0.056 |
| Mild, n (%) | 92 (68.15) | 33 (73.33) | 30 (66.67) | 29 (64.44) | |
| Moderate, n (%) | 15 (11.11) | 4 (8.89) | 9 (20.00) | 2 (4.44) | |
| **Myalgia, n (%)** | 69 (51.11) | 24 (53.33) | 24 (53.33) | 21 (46.67) | 0.745 |
| Mild, n (%) | 60 (44.44) | 21 (46.67) | 22 (48.89) | 17 (37.78) | |
| Moderate, n (%) | 8 (5.93) | 2 (4.44) | 2 (4.44) | 4 (8.89) | |
| **Fatigue, n (%)** | 57 (42.22) | 20 (44.44) | 22 (48.89) | 15 (33.33) | 0.306 |
| Mild, n (%) | 57 (42.22) | 20 (44.44) | 22 (48.89) | 15 (33.33) | |
| Moderate, n (%) | 0 (0.00) | 0 (0.00) | 0 (0.00) | 0 (0.00) | |
| **Headache, n (%)** | 63 (46.67) | 19 (42.22) | 26 (57.78) | 18 (40.00) | 0.109 |
| Mild, n (%) | 50 (37.04) | 15 (33.33) | 18 (40.00) | 17 (37.78) | |
| Moderate, n (%) | 13 (9.63) | 4 (8.89) | 8 (17.78) | 1 (2.22) | |
| **Fever, n (%)** | 3 (2.22) | 1 (2.22) | 1 (2.22) | 1 (2.22) | 0.365 |
| Mild, n (%) | 1 (0.74) | 1 (2.22) | 0 (0.00) | 0 (0.00) | |
| Moderate, n (%) | 2 (1.48) | 0 (0.00) | 1 (2.22) | 1 (2.22) | |
| **Flu-like, n (%)** | 1 (0.74) | 0 (0.00) | 0 (0.00) | 1 (2.22) | 0.365 |
| Mild, n (%) | 1 (0.74) | 0 (0.00) | 0 (0.00) | 1 (2.22) | |
| Moderate, n (%) | 0 (0.00) | 0 (0.00) | 0 (0.00) | 0 (0.00) | |
| **Somnolence**, n (%) | 7 (5.19) | 3 (6.67) | 3 (6.67) | 1 (2.22) | 0.547 |
| Mild, n (%) | 7 (5.19) | 3 (6.67) | 3 (6.67) | 1 (2.22) | |
| Moderate, n (%) | 0 (0.00) | 0 (0.00) | 0 (0.00) | 0 (0.00) | |
| **Diarrhea, n (%)** | 8 (5.93) | 4 (8.89) | 3 (6.67) | 1 (2.22) | 0.395 |
| Mild, n (%) | 8 (5.93) | 4 (8.89) | 3 (6.67) | 1 (2.22) | |
| Moderate, n (%) | 0 (0.00) | 0 (0.00) | 0 (0.00) | 0 (0.00) | |
| **Nausea, n (%)** | 15 (11.11) | 5 (11.11) | 8 (17.78) | 2 (4.44) | 0.132 |
| Mild, n (%) | 15 (11.11) | 5 (11.11) | 8 (17.78) | 2 (4.44) | |
| Moderate, n (%) | 0 (0.00) | 0 (0.00) | 0 (0.00) | 0 (0.00) | |

| **Adverse events** | | **Type of vaccines** | | | ***p*-value** |
|---|---|---|---|---|---|
| **Vomit, n (%)** | 1 (0.74) | 1 (2.22) | 0 (0.00) | 0 (0.00) | 0.365 |
| Mild, n (%) | 1 (0.74) | 1 (2.22) | 0 (0.00) | 0 (0.00) | |
| Moderate, n (%) | 0 (0.00) | 0 (0.00) | 0 (0.00) | 0 (0.00) | |
