## Supplementary figures and images for "Evaluation of the safety and immunogenicity of fractional intradermal COVID-19 vaccines as a booster: A pilot study"

### Supplementary Figure 1

**Supplementary Figure 1.**

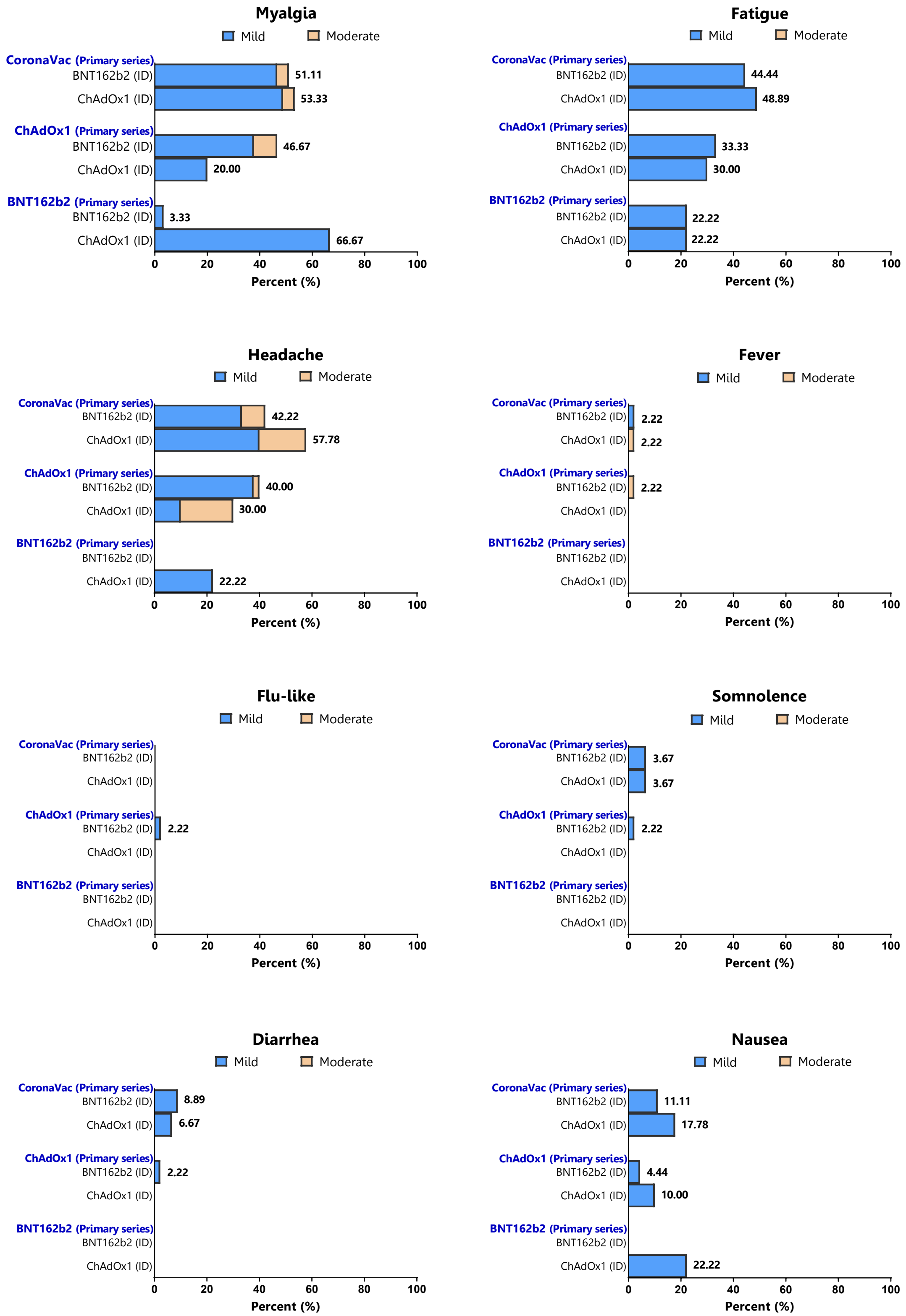

### Supplementary Figure 2

**Supplementary Figure 2.**

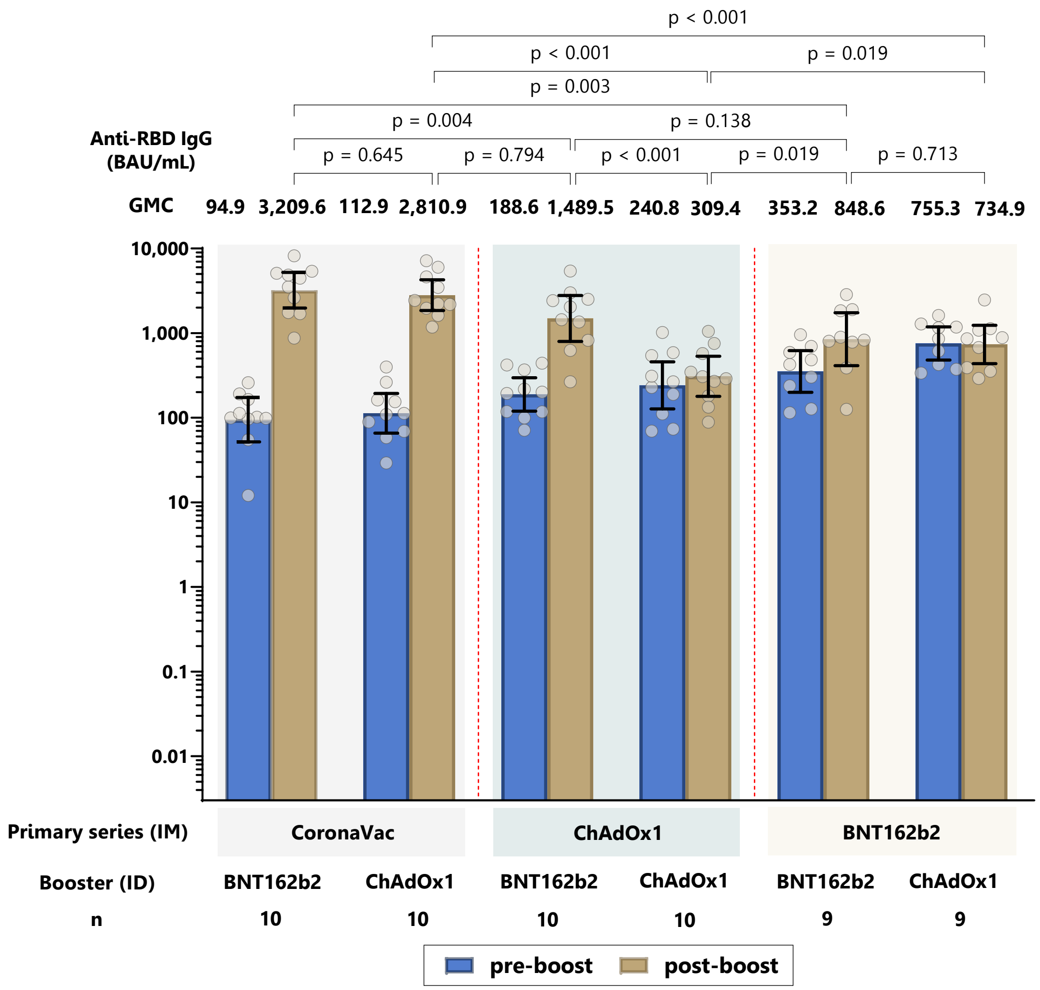
